# Estimates of the global burden of Japanese Encephalitis and the impact of vaccination from 2000-2015

**DOI:** 10.1101/19006940

**Authors:** Tran Minh Quan, Tran Thi Nhu Thao, Nguyen Manh Duy, Tran Minh Nhat, Hannah E. Clapham

## Abstract

Japanese encephalitis (JE) is a mosquito-borne disease, known for its high death and disability rate among symptomatic cases. Many effective vaccines are available for JE, and the use of a recently developed and inexpensive vaccine has been increasing over the recent years particularly with Gavi support. Estimates of the local burden and the past impact of vaccination are therefore increasingly needed, but difficult due to the limitations of JE surveillance. In this study, we implemented a mathematical modelling method combined with age-stratifed case data which can overcome some of these limitations. We estimate in 2015 JE infections caused 100,308 cases (95%CI: 61,720 - 157,522) and 25,125 deaths (95%CI: 14,550 - 46,031), and that between 2000 and 2015 307,774 JE cases (95%CI: 167,442- 509,583) were averted due to vaccination. Our results highlight areas that could have the greatest benefit from starting vaccination or from scaling up existing programs and will be of use to support local and international policymakers in making vaccine allocation decisions.

## Introduction

Japanese encephalitis (JE) is caused by Japanese encephalitis virus (JEV) – an arbovirus that belongs to the flavivirus genus, family flaviviridae. The main mosquito vectors are the Culex, especially Culex tritaenuirhynchus, which thrive in rice-paddy fields (Buescher and Scherer 1959; Self et al. 1973). JEV has a wide range of vertebrate hosts, noticeably the amplifying hosts are thought to be pigs and wading birds (SAGE Working Group on Japanese encephalitis vaccines 2014). Humans are dead-end hosts as viremia is not believed to reach levels that are infectious to mosquitoes (SAGE Working Group on Japanese encephalitis vaccines 2014). Only 1 in 25 to 1 in 1000 infections result in symptoms (Vaughn and Hoke 1992; SAGE Working Group on Japanese encephalitis vaccines 2014). However, the mortality rate of symptomatic cases is high - around 20-30% (Fischer et al. 2008), and around 30-50% of survivors experience significant neurological and psychiatric sequelae (Fischer et al. 2008).

The first JE case was documented in Japan in 1871 (WHO 2015). In 1924, a first JE outbreak in Japan caused more than 6, 000 cases and 3, 000 deaths in 6 weeks (Solomon 2006). Several outbreaks occurred subsequently in Asia (Hullinghorst 1951; Erlanger et al. 2009; Barzaga 1990). More recently, in 2005 large outbreaks occurred in northern India and Nepal, with 5,000 cases and 1,300 deaths (Solomon 2006). Currently, 24 Asia-Pacific countries are thought to be endemic for JE, with 3 billion individuals at risk of infection (WHO 2015).

The first vaccination was an inactivated mouse brain vaccine produced in Japan, used worldwide for 50 years. Although vaccine production halted in 2006, similar inactivated mouse brain vaccines are still produced locally in South Korea, Taiwan, Thailand and Vietnam (Yun and Lee 2013). The next vaccination, an inactivated a Vero cell vaccine (SAGE Working Group on Japanese encephalitis vaccines 2014), has been gradually replaced (since 1988) by a live attenuated vaccine (SA 14-14-2) produced in China. SA 14-14-2 is now widely used in Asia and funded by Gavi, greatly increasing the use. This vaccine requires only a single dose, is cheap to produce, and is safer than the mouse brain vaccine (SAGE Working Group on Japanese encephalitis vaccines 2014). In addition, a live attenuated chimeric vaccine was first licensed in Australia in 2012 (SAGE Working Group on Japanese encephalitis vaccines 2014).

WHO recommends two JE surveillance systems, i) a subnational system with sentinel hospitals, or ii) case-based nationwide surveillance. Each country implements one of these systems depending on available resources (Hills et al. 2009). WHO recommends diagnosis using JEV-specific IgM antibody-capture enzyme-linked immunosorbent assay (MAC-ELISA) in CSF at two time points (Donadeu et al. 2009; Burke and Leake 1988). Serum samples can be used, but false positives may result from cross-reactivity with other flaviviruses or vaccination (Solomon et al. 1998; Hills et al. 2009). Other tests that can confirm JE are plaque reduction neutralizing (PRNT), haemagglutination inhibition (HI), immunohistochemistry or immunofluorescence assay, reverse transcription polymerase chain reaction (RT-PCR) or virus isolation (Hills et al. 2009), though these are not often used.

The previous estimate of annual global JE cases was 67,900 with 13,600 – 20,400 deaths (Campbell et al. 2011). For this estimate a systematic review in 2011 collated case incidence data from endemic JE countries. Countries were then stratified into 10 incidence groups based on geographic, ecological and vaccine program similarities. The systematic review resulted in 12 key studies, which were then used to infer the incidence rate (IR) of the 10 incidence groups. However the estimation had some limitations; the surveillance quality of the 12 key studies varied and as the case incidence rate combines both the infection rate and vaccination, it is not possible to estimate the impact of vaccination.

Poor clinical outcomes and lack of specific treatment makes JE prevention a priority. Vaccination is the most effective method of prevention, however it is difficult to decide where vaccination should be implemented or to estimate the quantitative impact of vaccination (Fischer et al. 2008). In Nepal, one study estimated 3,011 JE cases were prevented in vaccinated districts from 2006 to 2012 (Upreti et al. 2017). Another study in Sarawak Malaysia estimated a 61% reduction in JE cases after the vaccination program, where climate effects were not taken into account, and 45% when the effects of climate were included (Impoinvil et al. 2013). The methods used in both these papers require good surveillance data before and after vaccination, which, though data is improving, is currently not widely available. Hence, new approaches are needed to estimate burden and vaccine impact.

In this study, we provide updated global JE burden and vaccination impact estimates using a modelling method which helps overcome some of the limitations of sparse and variable surveillance data. In addition, by simulating the model with and without the undertaken vaccination programs we are able to estimate the impact of vaccination on the number of global JE cases to date and identify areas that would benefit most from future vaccination.

## Results

There are two main stages to our analysis, summarized in flowcharts in Fig 1. In the first stage, we conducted a systematic review to collate age-stratified case data and a literature review to obtain vaccination information. We then fit a model to this data to estimate the transmission intensity or force of infection (FOI) for each study. In the second stage, we extrapolated the FOI for all endemic areas from our previous estimates. Using the processed population and vaccination data in all endemic areas, we used the model to generate burden quantities (cases) in two scenarios, with or without the JE vaccination programs that have been implemented.

**Figure 1.**
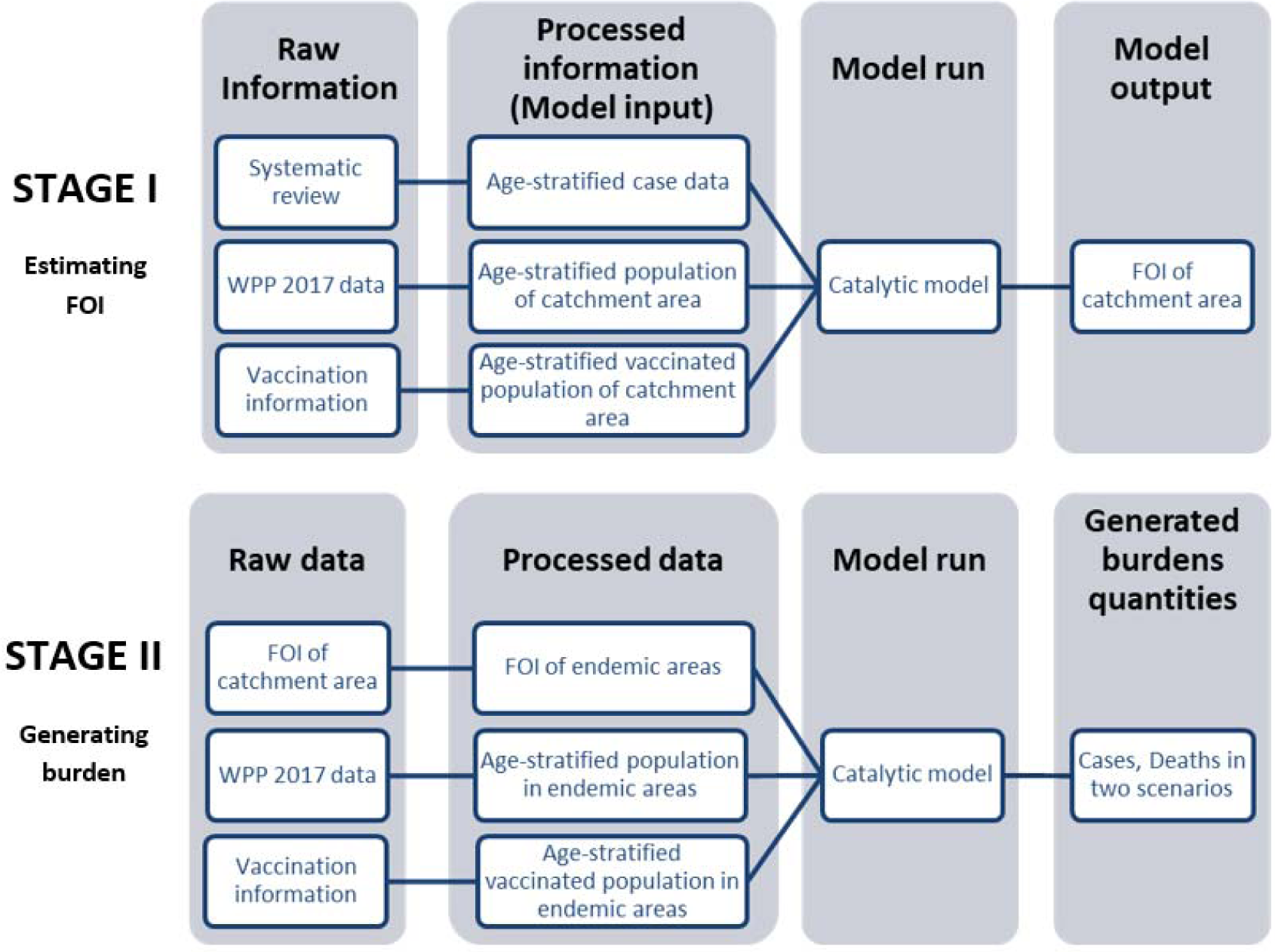
Flowchart describes two main stages in our analysis: Estimating FOI and generating burden. In Stage I we estimate FOI of all studies’ catchment area. In Stage II we then used the FOI estimates to generate global burden. Abbreviation: WPP: World Population Prospects

### Systematic review

A systematic review on October 11^th^ 2017 yielded 2337 initial results (Fig 2). 407 relevant studies were obtained after eliminating 1931 irrelevant titles and abstracts that were about molecular biology, policy, entomology, hosts other than humans, or were review papers. The obtained studies mainly comprised of reports of JE surveillance or epidemiological studies in one specific location. We also included modelling, economic evaluation or vaccine program assessment studies for possible eligible data sources in the references. We retrieved and read 261 full-text papers. Most of papers that we could not access were either old or not in English. In the systematic review process, a further 4 eligible studies were retrieved from references. 202 papers were then excluded as they did not contain age-stratified case data, and other 14 papers were also excluded because they had limited samples (less than 15 cases) or the study’ catchment area was not clear. Another 4 datasets from JE national reports were collated from Taiwan, Japan, and Sri Lanka. Finally, we had 53 studies that contained age-stratified case data (Fig 2). 42 of the 53 studies (79%) contained data from after 2000 only, 7 from before 2000 only and 3 from both time periods (Fig 2-Supp 1). 34 studies (64%) had data from 1-4 year time periods, 6 studies had data for between 5 and 9 years, and 11 studies had data for more than 10 years. The majority of the studies used the WHO JE case definition: JE IgM antibody in CSF or serum as confirmed by MAC-ELISA on patients with acute encephalitis syndrome. In the majority of studies patients were recruited from a sentinel hospital surveillance system, though these ranged in size from one to several hospitals. For studies with a consistent catchment area but for which data was collected in multiple years, we aggregated the age-stratified case data across years. Further details of the selected studies and data, including about catchment areas, sample collection methods, and vaccination programs are in Figure 2-Supp 1.

**Figure 2.**
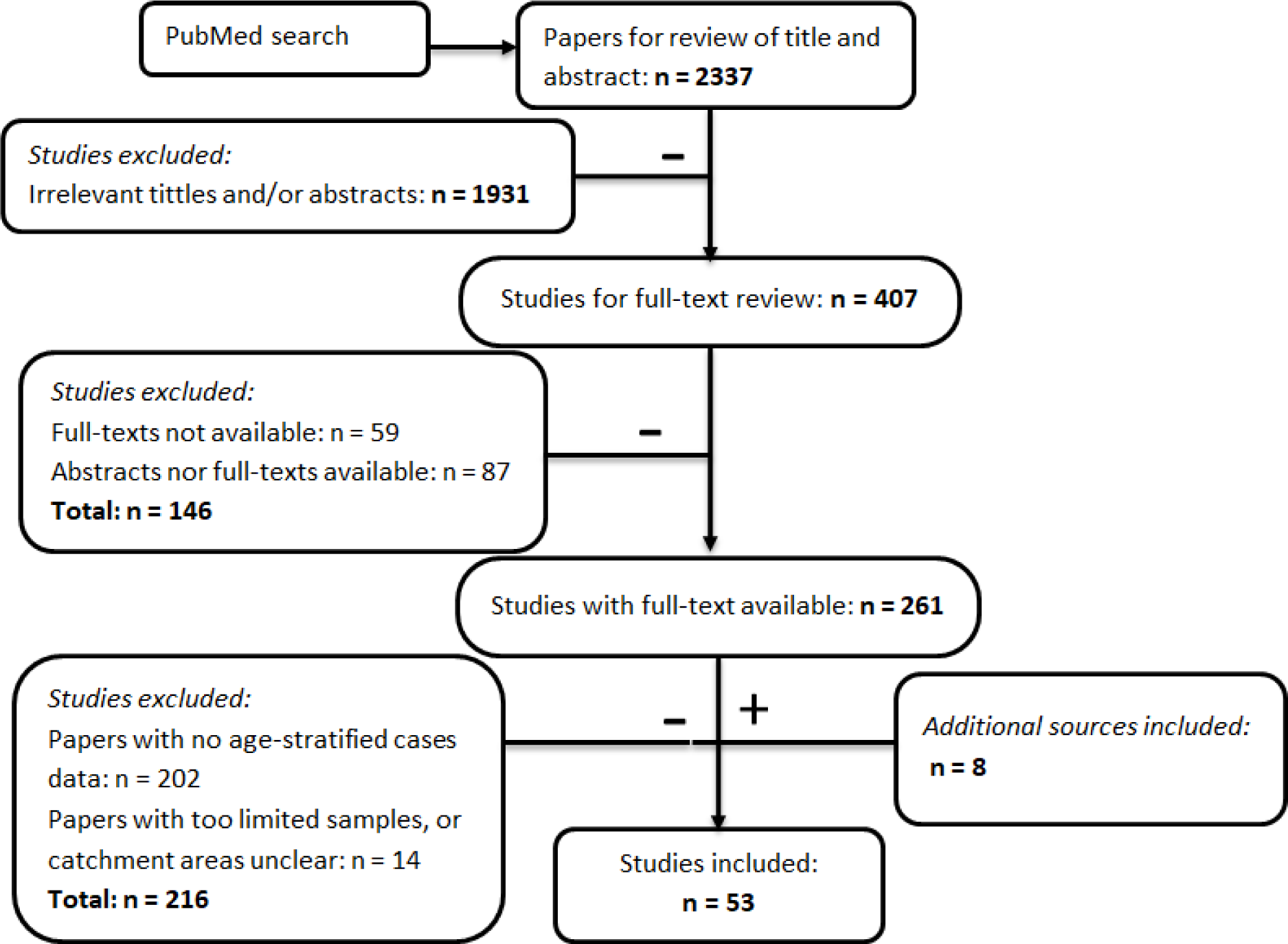
Flowchart describing the systematic review procedure searching for Japanese encephalitis age-stratified case data.

We obtained the vaccination information from three main sources: literature review, WHO, and Gavi (S2 Table). Campaign vaccination information was mainly from Gavi and routine vaccination was from WHO, while the literature contains both. When there were disagreements between the different vaccination information sources, we chose to use the information from the literature review. The total vaccinated population in each country using information obtained from this data from 2000-2015 is shown in Fig 3.

**Figure 3.**
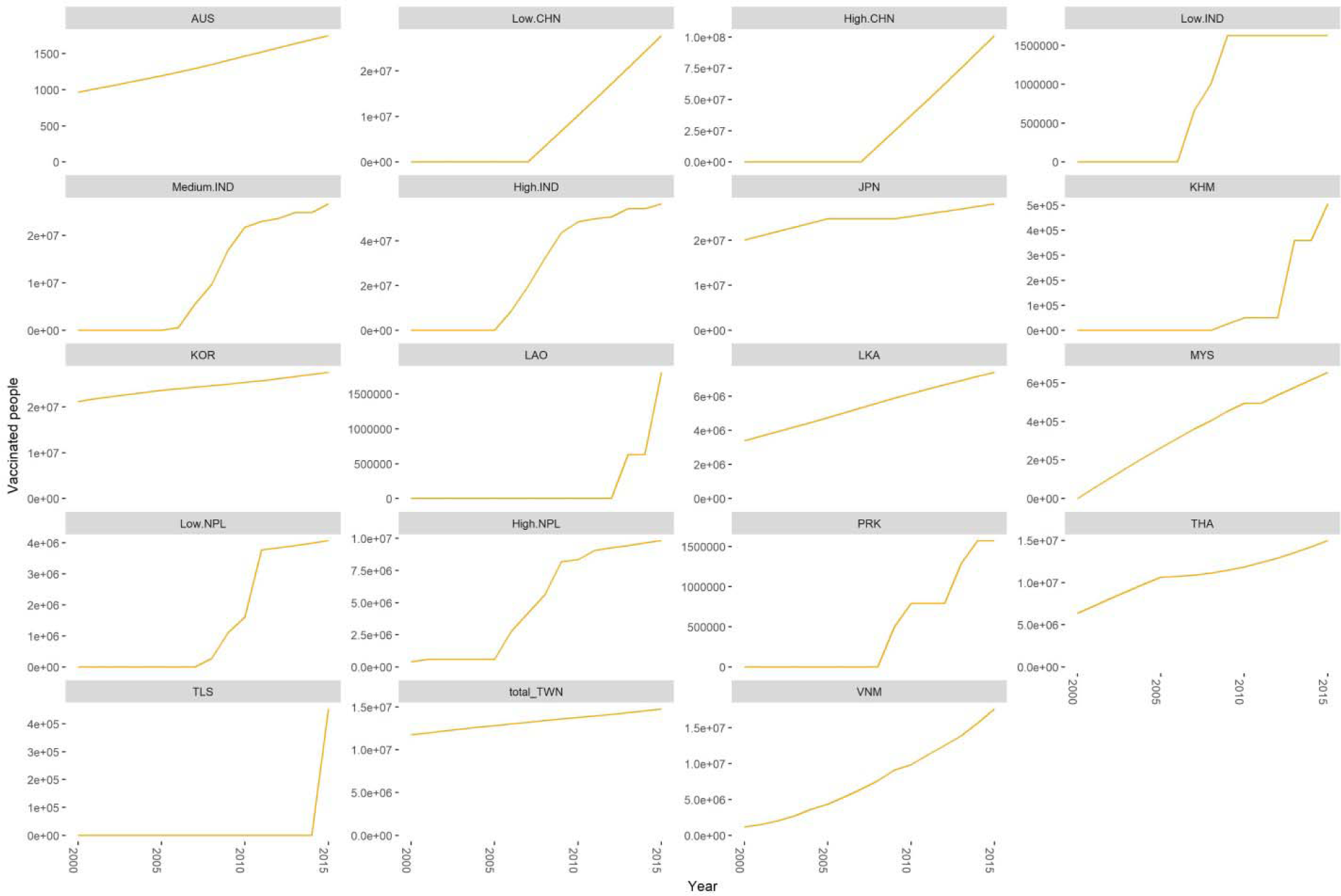
Estimated number of vaccinated individuals by region from 2000-2015. Abbreviation: BGD: Bangladesh, CHN: China, IDN: Indonesia, IND: India, JPN: Japan, KHM: Cambodia, KOR: South Korea, LAO: Laos, LKA: Sri Lanka, MMR: Myanmar, NPL: Nepal, PHL: Philippines, THA: Thailand, TWN: Taiwan

### Force of infection estimation from collated age-stratified data

From 53 studies, we made FOI estimates using the catalytic model from 53 unique catchment areas in 15 countries (Fig 4). All models converged well and mostly fit well to the data (Fig 4-Supp1). Our FOI estimates varied from 0.001 (95% CI: 0.000 - 0.002) in Japan to 0.507 (95% CI 0.419 - 0.582) in Guigang in China. Besides those extreme values, FOI were generally between 0.05 and 0.2, with a median of 0.09 (Fig 4). We also observed a wide variation in estimated reporting rates *ρ* between studies (S2 Fig). We estimated that the proportion of the population in study *k* and age group *i* that remained susceptible after vaccination *s*_*k,i*_, was different to the prior collated vaccination information in areas such as China, India, Japan, and Nepal (Fig 2-Supp 4).

**Figure 4.**
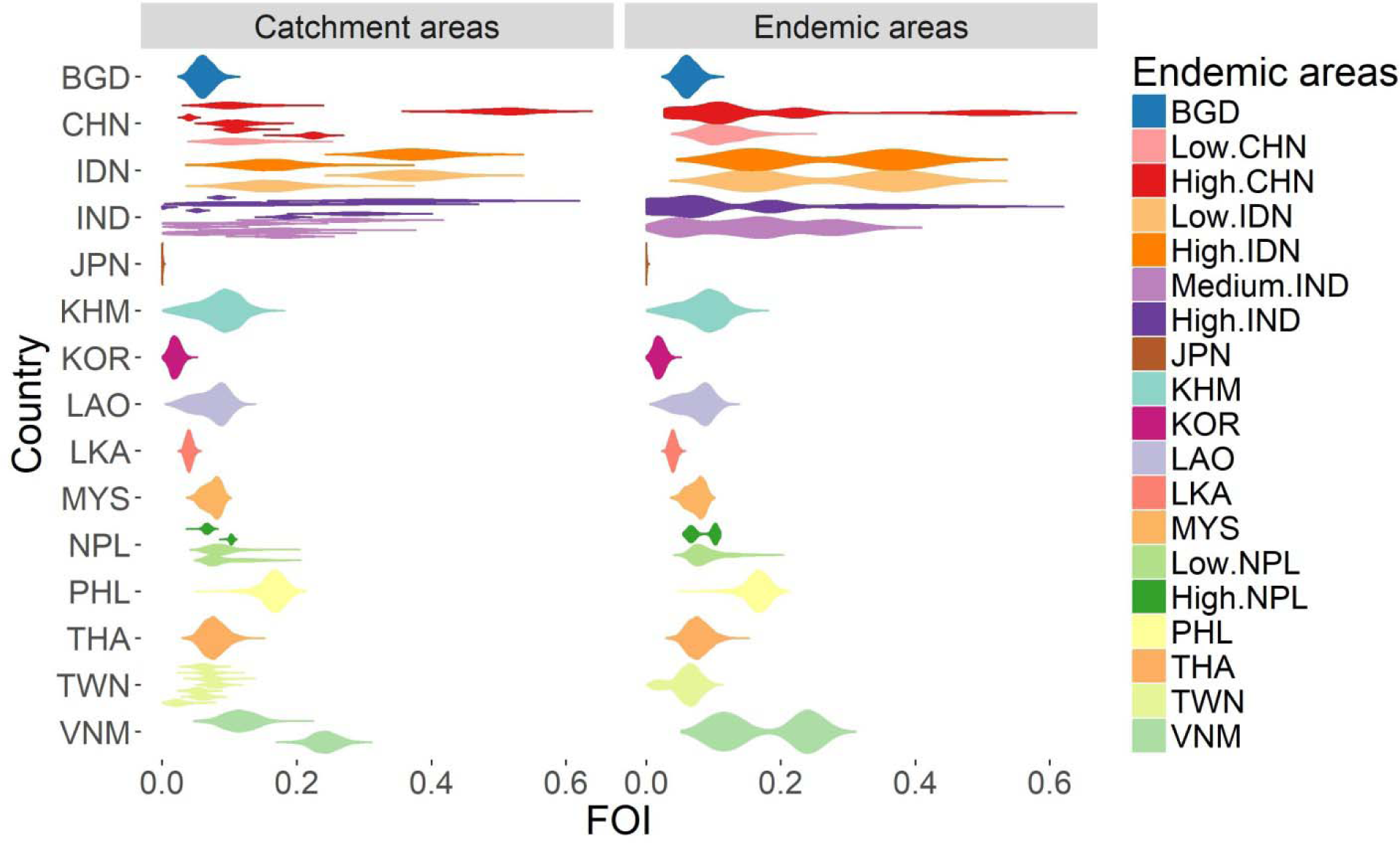
FOI distribution estimated from all studies’ catchment areas (on the left), which were used to infer the FOI distribution in all endemic areas (on the right). The colors are coded after the endemic areas as in the legend. Abbreviation: BGD: Bangladesh, CHN: China, IDN: Indonesia, IND: India, JPN: Japan, KHM: Cambodia, KOR: South Korea, LAO: Laos, LKA: Sri Lanka, MMR: Myanmar, NPL: Nepal, PHL: Philippines, THA: Thailand, TWN: Taiwan

### Inference of force of infection for all endemic areas

Based on the rules as mentioned above, we are able to infer FOI from available data for 24 endemic areas (Fig 2-Supp 4 and Fig 4). There were no studies in incidence group B (Australia, low incidence area in India, Pakistan, North Korea, Russia and Singapore). Since this group contains extremely low incidence areas, the FOI was assumed to have a lognormal distribution *1n*(*X*)*N*(0.01,1). For Indonesia, since the collated data was combined from various provinces across both the low and high incidence areas, we assumed the FOI to be the same in both areas.

### Burden and Vaccine Impact estimation

We estimate that from 2000 to 2015, there were 1,976,238 (95% CIs: 1,722,533 - 2,725,647) JE cases globally. By including known annual vaccination information in the catalytic model we estimate that in the same period had there been no vaccination there would have been 2,284,012 (95% CIs: 1,495,964 - 3,102,542) JE cases. Therefore we estimate that vaccination programs have prevented 307,774 JE cases globally (95% CI: 167,442-509,583) from 2000 to 2015 and vaccination programs similarly prevented 74,769 deaths from JE (95% CIs: 37,837-129,028). We estimate the greatest impact of vaccination from 2005 - 2010 due to large increase in vaccination in China in this time, and the impact of vaccination became more obvious over time (Fig 5). In 2015, we estimate vaccination reduced the number of cases globally by around 45,000 (from 145,542 (95% CI: 96,667 - 195,639) to 100,308 (95% CI: 61,720 - 157,522) (Fig 5).

**Figure 5.**
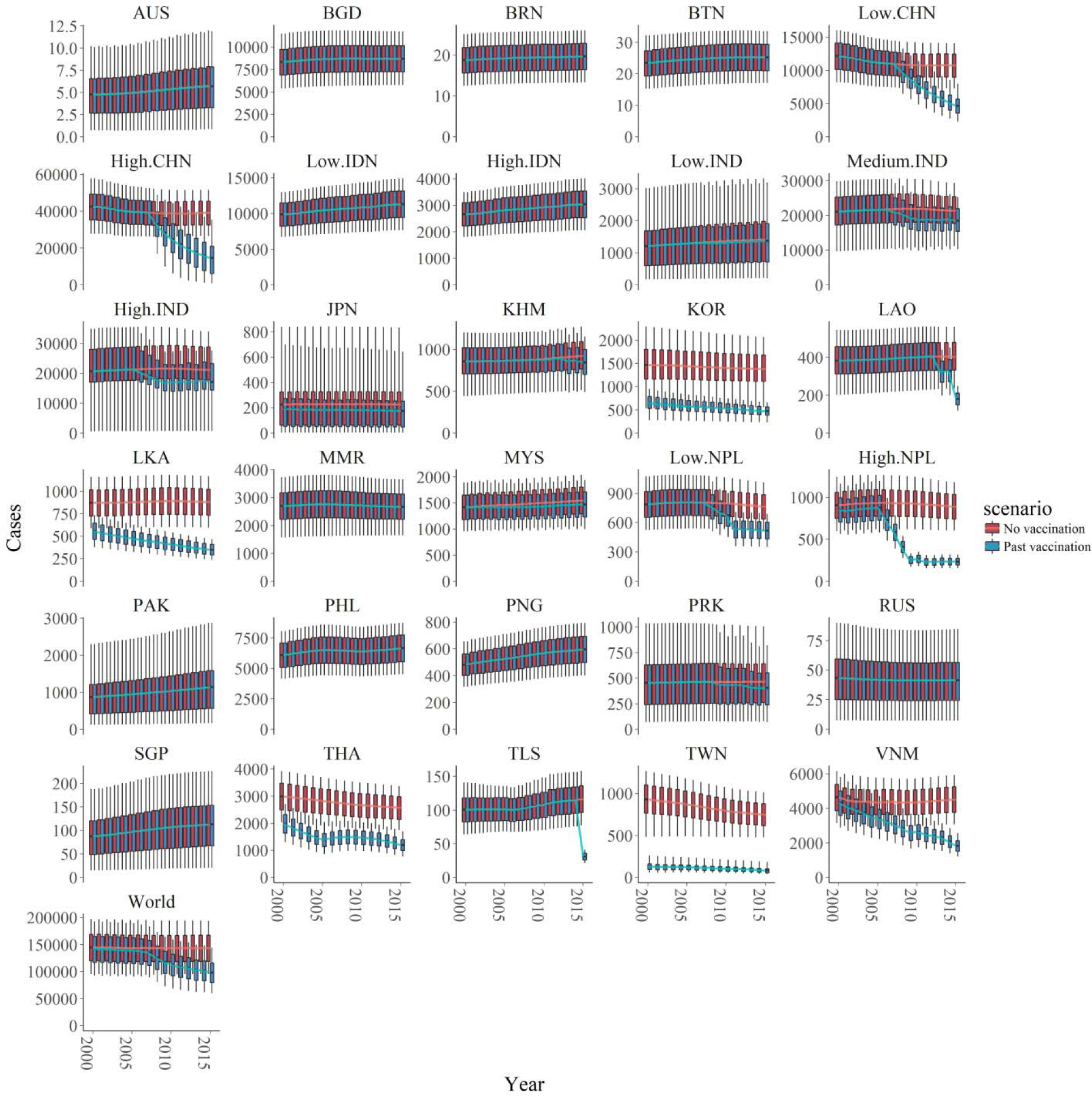
Number of estimated cases with and without vaccination of the 30 endemic areas and of the world from 2000 to 2015. The two scenarios, with or without vaccination, are also shown in blue and red respectively. In all areas, the boxplots represent the estimated cases with 95% credible intervals (also shown 1st quartile, 3rd quartile) with the solid lines showing the mean value of each interval. Abbreviation: AUS: Australia, BGD: Bangladesh, BRN: Brunei, BTN: Bhutan, CHN: China, IDN: Indonesia, IND: India, JPN: Japan, KHM: Cambodia, KOR: South Korea, LAO: Laos, LKA: Sri Lanka, MMR: Myanmar, MYS: Malaysia, NPL: Nepal, PAK: Pakistan, PHL: Philippines, PNG: Papua New Guinea, PRK: North Korea, RUS: Russia, SGP: Singapore, THA: Thailand, TLS: Timor-Leste, TWN: Taiwan, VNM: Vietnam

We estimated the highest number of cases in the high endemic area of China (around 40,000 annual cases in the no vaccination scenario and around 20,000 annual cases in vaccination scenario) and medium or high endemic areas in India (around 20,000 annual cases in no vaccination scenario and 15,000 annual cases in vaccination scenario for each area in recent years). On the contrary, areas like Australia, Brunei, Bhutan and Russia were estimated to have less than 100 annual cases with or without vaccination (Fig 5, Fig 6). All visualized burden estimates for every years and areas can be found in our interactive map (Duy 2018).

**Figure 6.**
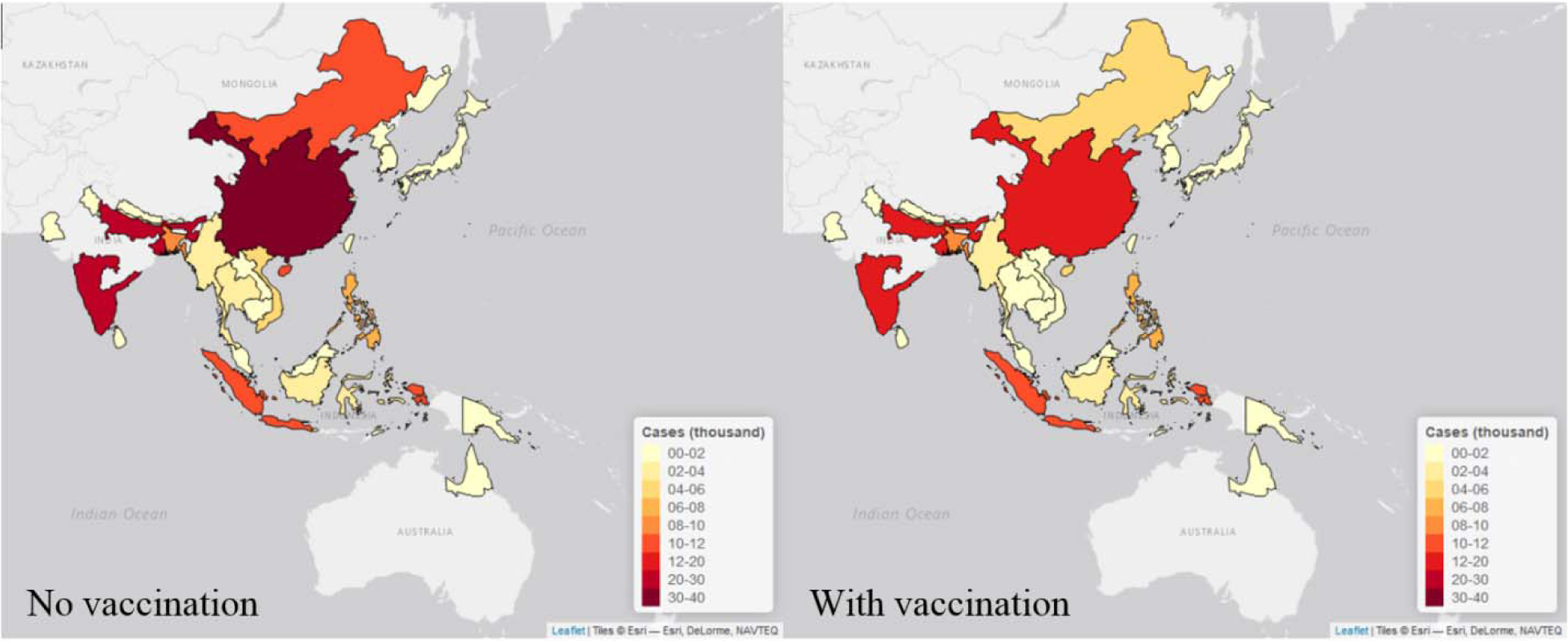
Maps of estimated cases (in thousand) in 30 endemic areas for two scenarios in 2015. Each endemic area is shaded in proportion to the area’s estimated cases in thousand as seen in the legend, with yellow shade is the lowest value and red shade is the highest value. The map on the left is the estimates from no vaccination scenario, and the right is from the vaccination scenario. The maps were made by *leaflet* package in R (Joe, Bhaskar, and Yihui 2017).

Vaccination impact can be observed in 19 areas where vaccination has been used (Fig 5). In areas like the low and high endemic area in China, medium and high endemic area in India, Cambodia, Laos, Nepal, North Korea, and East Timor though vaccination started recently, we estimate that the programs have achieved significant cases averted. Indeed, in the high endemic area in China, the routine vaccination programs only started in 2008 but contributed the most to the global cases reduction, with around 20,000 cases averted in China in 2015. We also observed a clear difference in cases between vaccination and no vaccination scenario in areas with intensive vaccination program in the past such as South Korea, Sri Lanka, Thailand, Taiwan, and Vietnam. For Japan, Australia, and Malaysia, though vaccination began a long time ago, we estimated there has been minimal vaccine impact. From the data we collated, no vaccine programs had occurred in in Bangladesh, Brunei, Bhutan, the low and high endemic areas in Indonesia, Myanmar, Pakistan, Philippines, Papua New Guinea, Russia, or Singapore so vaccination has had no impact.

### Sensitivity analysis

To assess the impact of uncertainties in our data and assumptions we performed extensive sensitivity analyses. Sensitivity analyses were conducted for endemic areas with uncertain coverage data, where both national and subnational data were available (China, India and Nepal), or where we did not have any studies. The majority of the results showed minimal changes compared to our original estimates (Fig 5 Supp1A-J). Cases estimated from Taiwan subnational data were higher by about 200 to 400 cases before 2004 (Fig 5 Supp1C, D). In some areas, we observed significant differences in the estimated cases when the coverage was changed: when the vaccine coverage reduced by 10% and 30% in Sri Lanka or by 30% in Thailand and Taiwan, the mean values of estimated cases increase by around 40, 100, 300, and 220 respectively (Fig 5 Supp1G, H). However these changes account for a small fraction of our original global estimates. Sensitivity analysis varying the assumed 100% vaccine effectiveness to 90% and 70% showed global case estimates changed minimally with this assumption (Fig 5 Supp1K, L). In addition due to concerns about possible changes in FOI over times, we also tested our assumption of constant FOI by fitting multiple-year data to a time-dependent catalytic model. Overall, the annual FOI estimates are comparable with the constant FOI (Fig 5 Supp1M).

## Discussion

In this paper, we updated the JE burden estimates with a mathematical modelling method using data we collated from a systematic review. We estimated that in 2015 there were around 100,000 JE cases globally. In addition, we estimate that vaccination programs averted around 45,000 JE cases in 2015.

For Japanese encephalitis, since humans are dead-end hosts and therefore vaccination does not lead to herd immunity, the FOI we estimated represents the constant spread of the disease from the animal reservoirs to humans. This spread depends on epidemiological factors related to JE transmission such as climate, rural-urban, mosquito distribution (especially Culex tritaenuirhynchus), and pig and rice field distributions (Le Flohic et al. 2013). This explains why our estimated FOI varies widely. Looking crudely at the pig density (Gilbert et al. 2018) and a Culex tritaenuirhynchus probability map (Miller et al. 2012) there appears to be a broad correlation of these factors with our estimates. The high FOI estimated in the south of China, Vietnam, and Philippines is consistent with the high pig density and high probability of Culex in these areas. We also estimated high FOI in India and Indonesia; however these countries only have high probability of Culex but low pigs density. This suggests that other potential animal reservoirs may contribute to the transmission in these countries, likely the wading bird or even poultry, although current evidence is limited (Lord, Gurley, and Pulliam 2015). In Taiwan, South Korea, and Japan the current estimated FOI is lower compared to other areas, respectively 0.061 (95% CI 0.013 - 0.093), 0.041 (95% CI 0.026 - 0.057) and the lowest, 0.001 (95% CI 0.000 - 0.002), despite these areas having high probability of Culex mosquito and high pig density. These countries have had high JE burdens in the past but we do not estimate so currently. This could be due to lack of recent data, or perhaps suggests urbanization, which reduces the proximity of humans to pig farms and rice fields (where the mosquitoes thrive), may play an important role in lowering transmission. This could also be due to uncertainties in the long term vaccination information in these areas. Further work will use environmental covariates to gain estimates of FOI on a smaller spatial scale and over time. In addition, changes in these covariates into the future should be considered in estimates of the future vaccine impact.

A strength of our Bayesian approach was the possibility to include prior information on vaccination, but also assess whether this was consistent with the ages distribution of observed cases. For China and Japan we estimated lower susceptible proportions after vaccination in certain age groups compared to calculated proportions from the available data. This suggests that there are a large number of immunized people in certain age groups due to past vaccination for which we did not have information. In Nepal and India, we also observed differences between the data and estimated susceptible proportion after vaccination, though the vaccination information for these countries was more readily available. For India, this could be explained by both uncertainty in vaccine efficacy and vaccination coverage data. From 2006 to 2011, SA 14-14-2 vaccine was used in India for campaigns. Though the vaccine reported nearly 100% efficacy in vaccine trails, the efficacy in India was reported to be as low as 30% to 40% (Vashishtha and Ramachandran 2015). A previous evaluation of vaccine coverage also showed that the coverage data in India was lower than reported (Murhekar et al. 2017). Further studies are needed to explore whether there are different vaccine efficacies in different places.

Using the FOI from 30 endemic areas, we projected the regional and global JE burdens as well as the vaccine impact. By region, our burdens estimates are highest in China and India, which aligns with previous literature (Heffelfinger et al. 2017). Our global estimate of around 100,000 cases annually is about 1.5 times higher than the previous estimate of around 70,000 cases (Campbell et al. 2011). Similar patterns are seen for the comparison area by area, in which our estimates are either higher than or comparable to the previous estimates (Table 1). It is not surprising that our estimates are higher, since our method more robustly takes into account under-reporting and different surveillance quality. In addition, the numbers we reported here are time-dependent and not static because our estimates include population changes and the progression of vaccination programs over time.

**Table1:**
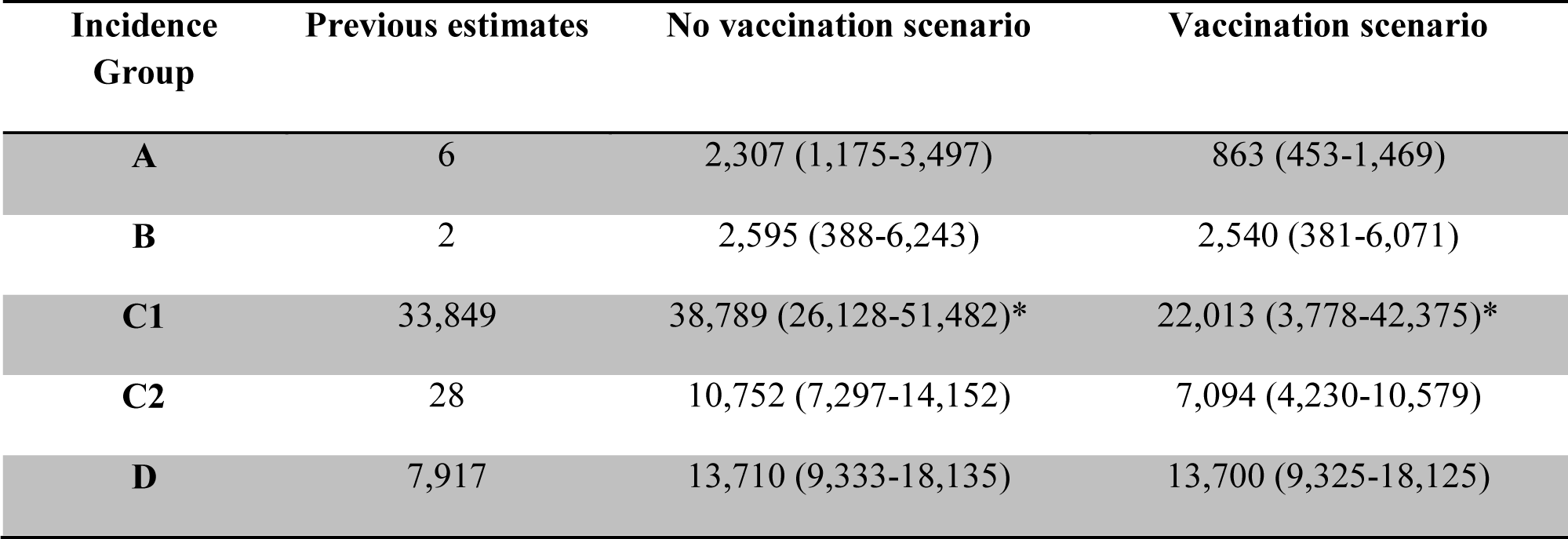
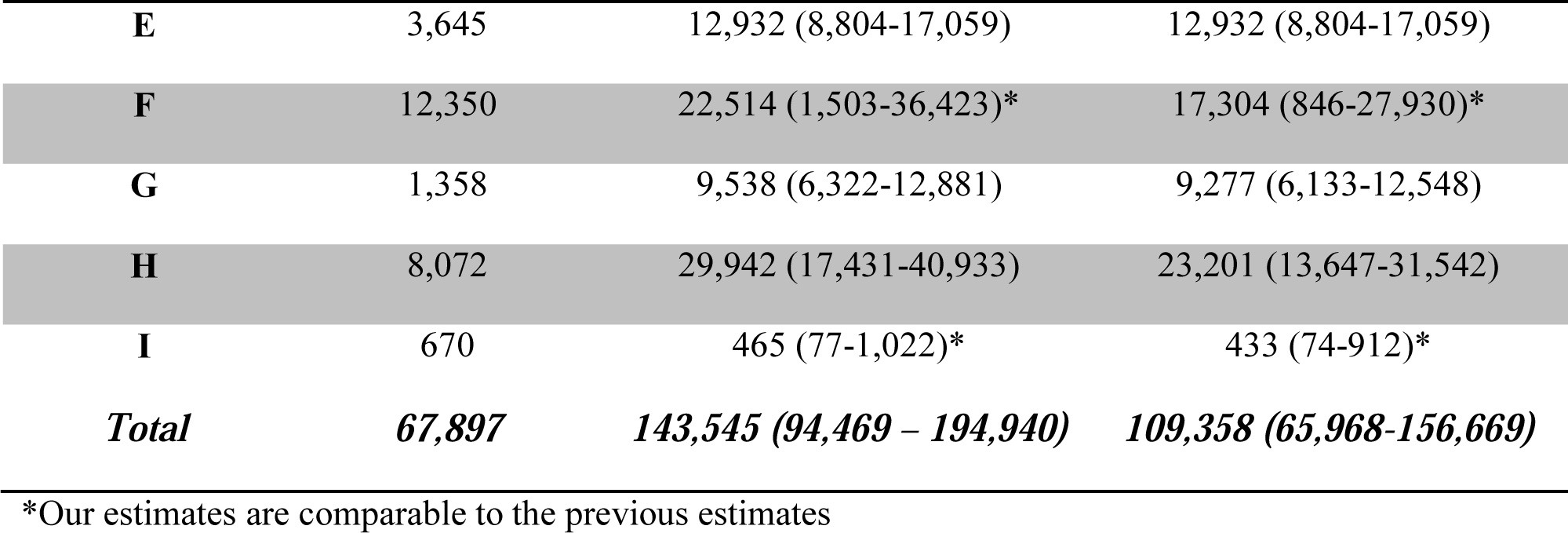
Comparing cases generated between previous estimates to our estimates. Group A: Taiwan, Japan, South Korea; Group B: Australia, low endemic area in India, Pakistan, Russia, Singapore; Group C1: high endemic area in China; Group C2: low endemic area in China; Group D: Cambodia, high endemic area in Indonesia, Laos, Sabah and Labuan in Malaysia, Myanmar, Philippines, Timor-Leste; Group E: low endemic area in Indonesia, Peninsular Malaysia, Papua New Guinea; Group F: high endemic area in India, high endemic area in Nepal; Group G: Bangladesh, Bhutan, Brunei, low endemic area in Nepal; Group H: Medium endemic area in India, Sarawak in Malaysia, Sri Lanka, Thailand, Vietnam; Group I: North Korea.

Though our methods are more robust, collating 53 studies (an additional 41 from the studies used in the previous burden estimate) (Campbell et al. 2011), and using age-stratified data to circumvent issues with reporting variation, there are still some limitations. As in the previous estimates of JE burden (Campbell et al. 2011), we made inferences for the whole country based on data from a few studies. However in our method we sampled from the FOI estimates from all studies to account for some of this uncertainty and variation. In addition, as in previous studies, a limitation is that we inferred the incidence metric (in our case, FOI) for areas without data, from FOI from other areas, based on previous classification of transmission in these countries. However our sensitivity analysis shows that this does not alter the global burden estimates greatly, though it may affect the country-specific burden estimates. (Campbell et al. 2011). Our future work incorporating the epidemiological factors into machine learning algorithms to extrapolate the FOI on smaller spatial scales will help in refining these estimates in the future.

We estimated only the impact of vaccination on cases from 2000 - 2015. Because the impact of vaccination will continue into the future as vaccinated individuals remain protected, our estimate will be an underestimate of the total impact of vaccination. In addition, our estimates will be an under-estimate of total vaccine impact as in some places vaccination programs have been running before 2000, and so vaccination had a large impact before 2000. However there is limited information in order to estimate transmission intensity before this time so we focused our work on 2000-2015. In this paper, we focused on cases (and to some extend deaths) from JE. However because a large number of cases have long-term sequelae after JE infection, focus just on case numbers does not describe fully the total burden of JE. Future work will refine the estimates of the proportion of individuals that die and that experience different long-term sequelae, to generate update our model to estimate JE Disability-Adjusted Life Year (DALY), particularly relevant for use in cost-effectiveness analyses for introduction of vaccination into new locations.

Since JE vaccination does not produce herd immunity, the transmission intensity can only be reduced by influencing the animal transmission cycle. Previous attempts to break the transmission cycle have been vector control and vaccination in pigs and wading birds, and this has been considered in modelling work (Khan et al. 2014). However they were either ineffective or up to now have been deemed economically and logistically intensive (Fischer et al. 2008). Further work considering pig vaccination in the context of these updated estimates of the burden of JE should be considered. We estimate that despite not interrupting transmission, human vaccination can be an effective strategy to reduce JE case numbers. This can be seen from the estimate that the majority of the reduction in global burden is due to the routine vaccination program in China from 2008. We estimate that India, East Timor, and Vietnam also have high transmission intensity, and residual cases despite vaccination, and therefore could further benefit from scaling-up the existing vaccination program. We estimated high transmission intensity in Indonesia, Papua New Guinea, and Philippines where there are no current vaccination programs, suggesting that vaccination in these areas should be a future priority. Future smaller scale estimates will support decisions on where within these countries could be best targeted for vaccination. For areas with a long history of JE vaccination such as South Korea, Sri Lanka, Thailand and Taiwan, (Fig 4), we estimate a substantial vaccine impact (Fig 5), though with cases still occurring. In other countries with a long vaccination history however, we estimate a minimal impact of vaccination (Fig 4 and 5), due to low estimated transmission intensity in Japan, low vaccine coverage in Malaysia, or both in Australia (though age-stratified data was not available in Australia). Our estimate of transmission intensity for Japan also has great uncertainty, as half the studies included data pre-2000 and we were able to find limited information on the long-running vaccination program there. Further work with serological data and further exploration of the drivers of JE transmission will help refine this estimate.

Assessing JE disease burden and vaccination program performance is important though difficult due of the lack solid surveillance programs worldwide. In our paper, we are able to estimate the disease burden and vaccine impact using a modelling method that is able to overcome some of the limitations of current surveillance. We estimate annually there are still 100,000 cases of this severe, but preventable disease, in Asia. The majority of remaining cases are focused in countries with still developing healthcare systems therefore vaccination should be a priority. The results generated from this study will help guide Gavi and other international and national public health agencies in deciding on when and where to direct their future investment into JE vaccination.

## Methods

### Systematic review

We performed a systematic review to find all available age-stratified case data for Japanese encephalitis in PubMed. We used the search terms “epidemiology” or “incidence” or “prevalence” or “public health” or “surveillance” or “distribution” in all fields with “Japanese encephalitis” in the title or abstract. All titles and abstracts were screened and we selected those in which the study contained age-stratified case data. We retrieved the full-texts for these selected abstracts and the abstracts were read by two independent reviewers to extract the age-stratified case data. From each study we also collected other information about the catchment areas, sample collection methods, diagnosis tests, and regional vaccination programs from the papers. A final consensus was reached for the final list of eligible full-texts. If abstracts were not available, the two independent individuals also tried to access and examine the full-texts. We also searched online for age-stratified case data from national JE surveillance reports.

We obtained vaccination information either from the study itself or from the literature review. Based on the review of JE vaccination programs reported from the World Health Organization (WHO) (Heffelfinger et al. 2017), we found that previous vaccination programs had occurred in 13 countries. We then undertook a literature search to find all vaccination information (target age group, vaccine coverage, types of vaccine used, years of vaccination) for these countries. We also collated historical routine vaccination program from country reported administrative doses data time series (from 2000 to 2015) compiled from WHO-UNICEF Joint Reporting (World Heath Organization 2018) and additional data from Gavi.

### Force of infection estimation

Force of infection (FOI) is the per capita rate at which susceptible individuals are infected by an infectious disease. In this study, we used a basic Muench’s catalytic model (Muench 1958) to estimate the constant age and time independent FOI using the case data we extracted during the systematic review process. A similar approach has been used to estimate the global dengue transmission intensity (Imai et al. 2016; Rodriguez-Barraquer, Salje, and Cummings 2019). As humans are dead-end hosts for JE, the FOI represents the FOI from the animal reservoir, and therefore is not impacted by human vaccination. This means vaccination can be included in the model simply as a removal of susceptible individuals by vaccination (or a reduction in risk of infection in this vaccinated group depending on vaccine efficacy) and will not alter the FOI. Therefore in this model, individuals can become immune to infection either by natural infection (depending on the force of infection) or vaccination.

To estimate the FOI (*λ*_*k*_), for each study *k*, taking into account vaccination and reporting rate for each study *k*, the modelled number of cases in a specific age group *i* is:

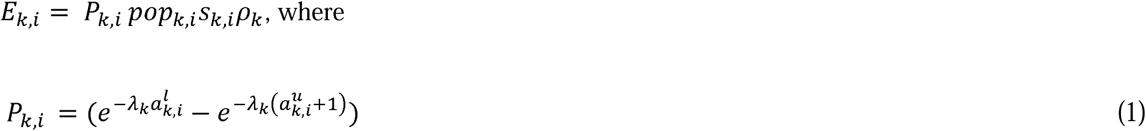

Where *P*_*k,i*_ estimates the incident rate of infection in each age group *i* (with lower and upper 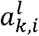 and 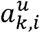 respectively), accounting for force of infection and susceptibility in that age group due to natural infection before this age. *pop*_*k,i*_ is the population size in each age group *i* of each study *k*, calculated from World Population Prospects 2017 data (United Nations-Department of Economic and Social Affairs-Population Division 2017). *s*_*k,i*_ is the estimated susceptible proportion in each age group *i* after vaccination for population in study *k*. The prior distribution of *λ*_*k*_ was an uninformative non-negative, normal distribution, *λ*_*k*_*norma1*(0,1000). To include the uncertainty in the vaccination information, we used an informative prior: 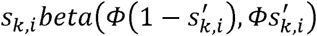, with 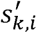 is the proportion of the population that remain susceptible after vaccination in age group *i* of study *k*, calculated from the vaccination information and the population demographics in the study’s catchment area. *Φ* represents the uncertainty of the vaccination information (we set *Φ* = 5).*ρ*_*k*_ is the reporting rate for each study, which is comprised of symptomatic rate and the reporting rate of the surveillance system and accounts for the different surveillance qualities of the different studies. Since *ρ*_*k*_ contains the symptomatic rate which reported to be less than 1% (SAGE Working Group on Japanese encephalitis vaccines 2014; Vaughn and Hoke 1992), we used an informative prior: *ρ*_*k*_*beta* (0.1,9.9).

The log-likelihood function for each study *k* is the sum of the multinomial log-likelihood and Poisson log-likelihood of total cases across all age groups.

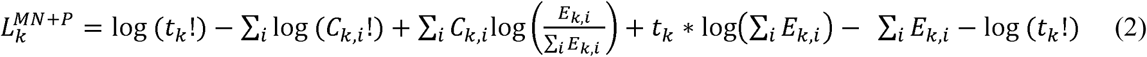

Where *t*_*k*_ is the total number of cases and *C*_*k,i*_ is the number of age-stratified cases in age group *i* in each study *k. E*_*k,i*_ is the modelled number of cases in a specific age group *i*.

For each dataset, we fitted the model in a Bayesian framework in RStan (Stan Development Team (2016)), estimating parameters *λ*_*k*_, *ρ*_*k*_,*s*_*k,i*_. The parameters *s*_*k,i*_, *ρ*_*k*_ were all estimated on a logit scale. We started 4 random chains, each with 16000 iterations and 50% burn-in period. Smaller step size of the Hamiltonian transition was manually set by increasing the adapt delta parameter in RStan to be 0.99. Model convergence was assessed visually.

We assumed that the JE vaccine has 100% effectiveness, which is reasonable given the reported high effectiveness of the vaccine ((WHO) 2012a, 2014, 2012b) and that the protection acquired from natural infection or vaccination was life-long. We further assumed the age distribution of the population within each country was homogenous across the country.

For our estimate, the endemic areas were defined to be the same as in the previous JE burden estimate (Campbell et al. 2011). For China, India, Nepal and Indonesia, where transmission intensity is diverse these countries were broken down to low, medium, or high endemic areas. In total, there are 30 endemic areas, spanning 24 countries. We inferred the FOI for each endemic area based on the FOI estimated from collated studies. The inference was based on two rules: 1) For each area, the FOI was obtained by sampling from the estimated FOI of all the studies that had catchment areas within that endemic area (if any). 2) For endemic areas in which no studies were conducted, the FOI was inferred to be equal to the FOI of the area in the same incidence group defined by (Campbell et al. 2011).

### Burden and vaccine impact estimation

Once the inferred FOIs for each endemic area were obtained, we generated the number of cases in each year *t* (from 2000 to 2015) in endemic area *d* for each age group *a* from 0 to 99 years old *t* and scenario *m* (described below) using the function (similar to the model used to estimate FOI (equation 1)):

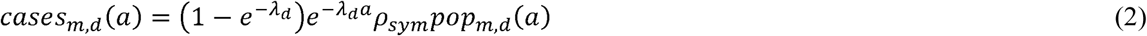

*λ*_*d*_ is the FOI of that area (assumed constant over time and age independent). The term 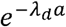 is the decrease in proportion of susceptible population due to natural infection. *ρ*_*sym*_ is symptomatic rate, sampled from *uniform* 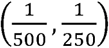(SAGE Working Group on Japanese encephalitis vaccines 2014). *pop*_*m,d*_(*a*) is the susceptible population of age *a* in endemic area *d* in year *t* under scenarios *m* and was interpolated from World Population Prospects 2017 data (United Nations-Department of Economic and Social Affairs-Population Division 2017). To assess the impact of previous vaccination programs, the population *pop*_*m,d,t*_(*a*) was different for each vaccination scenario *m*: with or without vaccination. The vaccination scenario used the collated information about past vaccination programs and assumed that the number of vaccinations given each year to each age meant that this number of the relevant age groups in the population were not susceptible to infection from this year onwards. This takes into account aging of the vaccinated population and any changes in the vaccination programs over time.

Although the mortality rate of JE varies, the reported ranges are from 20-30% (Fischer et al. 2008). We sampled the mortality rate from *uniform*(0.2,0.3) and multiplied it by the estimated number of *cases*_*m,d,t*_(*a*) to generate age-specific JE-induced deaths.

## Data Availability

This study conducted a literature review and collated all data on age-stratified JE cases from these papers. The full list of these papers and the
data extracted is available in the supplement.

## Supplementary Files Legends

**Figure 2 Supp 1: Studies from the systematic review that contain age-stratified case data**. Abbreviation: AUS: Australia, BGD: Bangladesh, BRN: Brunei, BTN: Bhutan, CHN: China, IDN: Indonesia, IND: India, JPN: Japan, KHM: Cambodia, KOR: South Korea, LAO: Laos, LKA: Sri Lanka, MMR: Myanmar, MYS: Malaysia, NPL: Nepal, PAK: Pakistan, PHL: Philippines, PNG: Papua New Guinea, PRK: North Korea, RUS: Russia, SGP: Singapore, THA: Thailand, TLS: Timor-Leste, TWN: Taiwan, VNM: Vietnam

**Figure 2 Supp 2: PRISMA 2009 flowchart**.

**Figure 2 Supp 3: PRISMA Checklist**.

**Figure 3-Source Data: Vaccine information and how it was used in our model**. Abbreviation: AUS: Australia, BGD: Bangladesh, BRN: Brunei, BTN: Bhutan, CHN: China, IDN: Indonesia, IND: India, JPN: Japan, KHM: Cambodia, KOR: South Korea, LAO: Laos, LKA: Sri Lanka, MMR: Myanmar, MYS: Malaysia, NPL: Nepal, PAK: Pakistan, PHL: Philippines, PNG: Papua New Guinea, PRK: North Korea, RUS: Russia, SGP: Singapore, THA: Thailand, TLS: Timor-Leste, TWN: Taiwan, VNM: Vietnam.

**Figure 4 Source data: Estimated FOI and studies used/assumptions of 30 endemic areas**.

**Figure 4 Supp 1: Model fit of all age-stratified case data**. For each study, the red dots with red vertical lines are the mean cases by age group estimated from the model with 95% credible interval. The blue dots are the cases by each age group.

**Figure 4 Supp 2: Estimated reporting rate from all studies**. For each study, the dots with vertical lines are the mean reporting rate estimated from the model with 95% credible interval. The colors represent the endemic areas as seen in the legend.

**Figure 4 Supp 3: Susceptible proportion after vaccination in study population**. For each study, the red dots with red vertical lines are the mean susceptible proportion after vaccination by age group estimated from the model with 95% credible interval. The blue dots with blue vertical lines are the mean susceptible proportion after vaccination by age group calculated from vaccination information with generated 95% credible interval from the beta distributions.

**Figure 5 Supp 1: Results of sensitivity analyses**

